## Supporting information for "An economic evaluation of *Wolbachia* deployments for dengue control in Vietnam"

**Project sites and stratification of dengue cases**

Settings were selected based on their share of the national dengue burden, considering both historical notified dengue case numbers and model-projected dengue case numbers to account for the known under ascertainment of disease burden in routine disease surveillance. Data on the number of dengue cases notified to the national dengue surveillance system by district each year 2016 – 2019, was provided by the Department of Preventive Medicine of the Vietnam Ministry of Health. The model-projected dengue case numbers were based on the projections by Bhatt *et al*. [1].

The total projected number of symptomatic dengue cases were broken down into the following severity categories;

1. Sought no formal treatment: self-managed cases disrupt the routine of the individual (e.g. not going to work or school) but do not result in seeking treatment at a formal private or public healthcare facility. Such cases may be untreated, self-treated (e.g. using medicines from a pharmacy) or treated in informal settings.
2. Outpatient cases: outpatient cases are severe enough for formal medical treatment to be sought but are managed on an outpatient basis, e.g. dengue (ambulatory) clinics.
3. Hospitalized cases: hospitalised cases are severe enough to require hospital admission and repeated observation by trained medical staff.
4. Fatal cases: fatal cases whereby acute dengue infection is the leading cause of death.

More general data and information regarding the characteristics/demographics of these study populations are available from the General Statistics Office of Vietnam [2].

### Health burden

*Burden of non-fatal cases*

The DALY burden of non-fatal cases was estimated using the overall disability weights from Zeng *et al*. [3] (Supporting Table S1). It was assumed that a self-managed case had the same disability weight as an outpatient case. These weights include persistent symptoms for which there is some uncertainty [4]. In the sensitivity analysis, we used the lower bound of the weights accounting only for the acute phase of illness.

| **Supporting Table S1: DALY disability weights (taken from [3])** | | |
| --- | --- | --- |
|  | **Median** | **95% Certainty interval** |
| Overall DALYs weight per episode (including persistent symptoms) | | |
| Outpatient episode | 0.0307 | 0.0170-0.0917 |
| Hospitalized episode | 0.0351 | 0.0241-0.0960 |
| DALYs per episode weight (acute phase only) | | |
| Outpatient episode | 0.0107 | 0.0057-0.0151 |
| Hospitalized episode | 0.0152 | 0.0100-0.0201 |

*Burden of fatal cases*

Based on GBD 2019 study we assumed a mortality rate of 11.65 per 100,000 symptomatic dengue episodes [5]. Based on the estimated number of deaths and years of life lost (YLL) from the GBD 2019 study we estimated that on average a fatal case corresponds to 55 years of life lost (undiscounted) [5].

**Economic burden**

Cost of illness data

The cost of illness data is summarised in Supporting Table S2. The cost of illness related to outpatient and hospitalized dengue patients was based on the data collected from Lee *et al*. [6]. There is little published data regarding the costs in Vietnam of informal medical care for dengue, such as the costs associated with patients treating themselves at home with medications obtained from pharmacies or traditional medicine practitioners. Shepard *et al*. [7] projected that the average cost of a self-managed dengue case (i.e. informal medical care) in Vietnam was approximately US$17.32 per case (adjusted to 2020 prices). The assumed cost of illness data for different types of dengue cases are summarised in Table 4. Based on Lee *et al.* [6] it was estimated that 55.3% of the direct medical costs of hospitalized and outpatient cases would be incurred by the healthcare providers and all other costs would be borne by the patients. The cost of illness related to fatal cases was estimated using the human capital approach – based on the number of the YLL of a fatal case up to the average retirement age of 61 and the GDP per capita (US$3,521[8]). The costs were discounted at 3% per year [9].

All of the costs were adjusted for inflation to 2020 prices using Vietnam specific GDP deflators [10, 11].

| **Supporting Table S2: Costs of illness for different dengue cases (2020 prices)** | | | |
| --- | --- | --- | --- |
| **Case type** | **Direct medical costs (95% CI)** | **Direct non-medical costs (95% CI)** | **Productivity costs (95% CI)** |
| Sought no formal treatment [7] | US$5.77  (4.60-7.09)^1^ | US$0  (0-0) | US$11.55  (8.41-15.79)^2^ |
| Sought formal treatment - outpatient case [6] | US$28.95  (24.49-33.40) | US$10.02  (7.79-14.47) | US$30.06  (22.27-42.31)^3^ |
| Sought formal treatment - hospitalized case [6] | US$92.41  (69.03-120.25) | US$56.78  (48.99-64.58) | US$74.60  (53.44-99.09)^3^ |
| Fatal cases | - | - | US$ 84,901.79 |
| *^1^ Range based on the uncertainty interval for the direct medical costs of the outpatient and hospitalized cases*  *^2^ Range based on the uncertainty interval for the productivity costs of the outpatient and hospitalized cases*  *^3^Also includes the losses of caregivers* | | | |

*Government’s current dengue prevention and control activities*

The costs related to the government’s current dengue prevention and control activities were obtained from Vietnam’s Ministry of Health records. Using the total budgets for the provincial governments for these activities in 2019 and 2020, we estimated a province specific average cost per person (based on the average annual total cost of the activities and the population size of the province) (Supporting Table S3). This was multiplied by the total population of the targeted setting to estimate a setting specific cost for these activities. We assumed that the costs related to these activities would decrease by 75% after the intervention become effective in line with the reduction in case numbers.

| **Supporting Table S3: Annual costs related to the government’s current dengue prevention and control activities** | | |
| --- | --- | --- |
| **Setting** | **Cost per person (US$)** | **Projected cost per year (US$)** |
| Hồ Chí Minh | 0.030 | 271,540 |
| Hà Nội | 0.040 | 320,480 |
| Đà Nẵng | 0.042 | 47,977 |
| Cần Thơ | 0.033 | 40,622 |
| Thuận An | 0.031 | 18,307 |
| Dĩ An | 0.031 | 14,575 |
| Thủ Dầu Một | 0.031 | 9,875 |
| Biên Hòa | 0.038 | 39,974 |
| Nha Trang | 0.065 | 27,275 |
| Vũng Tàu | 0.052 | 18,479 |

**Cost of the intervention**

The intervention was divided into the following phases:

- “Preparation” phase (1-year duration): includes establishing insectaries and a mosquito colony, laboratories, site offices, local regulatory approval, hiring staff, baseline entomological surveys (including insecticide resistance monitoring), and planning and administering the programme and pre-release community engagement.
- “Release” phase (6-month duration) involves the release of *Wolbachia* mosquitoes over target areas by applying the resources established during the preparation phase and monitoring the progress of *Wolbachia* introgression.
- “Short-term monitoring” phase (3-month duration), ongoing surveillance of the mosquito and human population is conducted in the release area and any requited rereleases are conducted.
- “Long-term monitoring” phase (10-year duration): includes reduced small ongoing costs associated with entomologic monitoring, surveillance and administration etc.

The costs for preparation, release and short-term monitoring phases were informed by the WMP accounts for the Thủ Dầu Một and Mỹ Tho project sites in Vietnam. The projected cost of future releases was adjusted to account for the fixed scaling of some cost types across multiple project sites and to reflect the realistic expectation that subsequent releases would be conducted with a faster timeline than the project sites. Based on WMP’s implementations in other countries the average undiscounted cost of the long-term monitoring phase was assumed to be US$0.20 per person covered – to account for a small ongoing cost associated with a low level of entomologic monitoring, surveillance and administration etc (Supporting Table S4).

Using this data from these two sites, we estimated an average cost per person covered across different phases of the intervention (Supporting Table S4). The average population density of these two sites was 12,411 inhabitants per km^2^). This was similar to the average population density of the investigated settings (14,625 inhabitants per km^2^) (Table 2).

All of the costs were adjusted for inflation to 2020 prices using Vietnam specific GDP deflators [10, 11].

Note that focusing on the cost per person would not necessarily be suitable if instead of looking at the cost-effectiveness of high burden cities we were also considering larger geographical areas or areas with a lower population density

| **Supporting Table S4: Projected undiscounted unit costs of the *Wolbachia* deployments (2020 US$ prices)** | | | | | |
| --- | --- | --- | --- | --- | --- |
|  | **Preparation phase**  **(12 months)** | **Release phase**  **(6 months)** | **Short-term monitoring phase (3 months)** | **Long-term monitoring phase – (10 years and 3 months)** | **Projected total** |
| **Average cost per person covered (US$)** | 3.97 | 3.23 | 1.32 | 0.20 | 8.72 |
| **Average Cost per km^2^ covered (US$)** | 49,260.39 | 40,147.57 | 16,382.35 | 4,889.31 | 108,265.15 |

**Further methodological details**

The area and population size of the proposed *Wolbachia* release areas were adjusted to account for the fact that releases would not cover a whole administrative area – as large public areas are excluded from releases and there is a minimum population density threshold below which releases would not be performed. Therefore, release area size and population values are smaller than the corresponding administrative district boundaries.

No specific health economic analysis plan was developed or previously published for this study. In addition, there were no approaches to engage patients, the general public, or stakeholders in the design of the study.

In terms of characterising heterogeneity and distributional effects, it was assumed that benefit of the intervention was equally experienced among the targeted populations. No specific adjustments were made to reflect priority populations. However, the impact within the different settings was investigated.

**Discussion of further limitations**

**The assumed costs of the intervention and economies of scale:** The same estimated cost per person covered was used for all of the project sites (based on data from the Thủ Dầu Một and Mỹ Tho project sites (Supporting Table S4)). However, in reality, the cost would via due to economies of scale and the population density of each setting. The population density of the Thủ Dầu Một and Mỹ Tho project sites was 12,411 inhabitants per km^2^. Note that because as the population density increases the cost per person covered decreases, the cost in areas with a higher population density would be overestimated– providing a conservative estimate. However, setting specific costs for some of the smaller locations with a lower population density could be underestimated.

**Government’s current dengue prevention and control activities:** The costs related to the government’s dengue prevention and control activities were obtained from Vietnam’s Ministry of Health records from 2019 and 2020 – estimating an average cost per person within the different provinces (based on the total cost of the activities and the population size of the province). However, in practice, the costs would not be uniformed distributed across a province and would be more likely to occur in urban areas. There would therefore be variation in the costs and the level of vector control activities across different settings that are not being captured. In addition, there is also uncertainty regarding how these activities would change once the *Wolbachia* deployments have reduced the burden of dengue. It should be noted that since 2021, as for many other infectious diseases, the provincial budget for dengue prevention and control no longer receives funding from the central government, known as the National Health - PopulationTarget Program. The provincial government is now fully responsible for allocating the budget for all the prevention and control activities in their province, with technical support from the national Department of Preventive Medicine.

**Cost of illness data**: The costs for the outpatient and hospitalized dengue cases related to a study conducted at Khánh Hòa General Hospital (Khánh Hòa province Oct 2011- Oct 2012 [6]. It was assumed it was generalisable to other settings in Vietnam, however, in practice, there will be a degree of variation in the cost of illness. The average cost of illness could also be influenced by the age distribution of the cases and therefore change over the course of the intervention.

**The type of model used to quantify the effects**: The static model we used did not account for the fact that targeting *Wolbachia* in a high-risk setting (particularly large cities) not only protects people who live in these areas, but also potentially protects visitors to these areas. If cities normally act as a source of infection that seeds outbreaks in local surrounding areas within Vietnam, surrounding areas may receive some indirect protection even if they don’t contain any *Wolbachia* mosquitoes. This would increase the public health impact. On the other hand, the static model used did not account for how the effectiveness of the *Wolbachia* deployments may change over time. For example, it is possible that as herd immunity levels decline over time (due to new births into the population) the effectiveness of this intervention would decrease. To account for this, we assumed a maximum reduction in the number of cases due to the intervention of 85% and only considered up to 25 years of benefit (with only 75% effectiveness and 20 years of benefit in the base case). Furthermore, the static model would not account for how the age distribution of cases changes as a result of the *Wolbachia* program. Decreasing the incidence of dengue could increase the average age of a secondary infection and change the age distribution of the cases. This will affect the number of DALYs averted for fatal cases. In addition, the model does not account for the spatial dynamics of the dengue transmission and how individuals could get infected when traveling to unprotected areas.

| **Supporting Table S5: Baseline epidemiology burden of dengue across the study settings** | | | | | | | | | | | | |
| --- | --- | --- | --- | --- | --- | --- | --- | --- | --- | --- | --- | --- |
| **Setting** | **Baseline annual epidemiology burden** | | | | | | **Baseline annual economic burden (2020 US$ prices)** | | | | | |
|  | **Sought no formal treatment** | **Sought formal treatment- outpatient** | **Sought formal treatment - hospitalized** | **Total number of cases** | **Fatal cases** | **Baseline DALYs burden** | **Cases that sought no formal treatment** | **Outpatient cases** | **Hospitalized cases** | **Fatal cases** | **Government’s current dengue prevention and control activities** | **Total economic burden** |
| Hồ Chí Minh | 103,574 | 37,649 | 20,359 | 161,582 | 18.8 | 6,085 | 1,793,724 | 2,598,869 | 4,556,207 | 1,598,047 | 315,602 | 10,818,388 |
| Hà Nội | 74,228 | 26,981 | 14,591 | 115,800 | 13.5 | 4,361 | 1,285,500 | 1,862,520 | 3,265,276 | 1,145,265 | 220,268 | 7,879,041 |
| Đà Nẵng | 7,903 | 2,873 | 1,554 | 12,329 | 1.4 | 464 | 136,870 | 198,306 | 347,661 | 121,939 | 37,387 | 852,752 |
| Cần Thơ | 12,413 | 4,512 | 2,440 | 19,366 | 2.3 | 729 | 214,980 | 311,478 | 546,067 | 191,528 | 34,964 | 1,304,675 |
| Thuận An | 8,401 | 3,054 | 1,651 | 13,106 | 1.5 | 494 | 145,485 | 210,789 | 369,545 | 129,614 | 18,280 | 873,741 |
| Dĩ An | 6,691 | 2,432 | 1,315 | 10,439 | 1.2 | 393 | 115,883 | 167,900 | 294,353 | 103,242 | 15,800 | 695,953 |
| Thủ Dầu Một | 4,994 | 1,815 | 982 | 7,791 | 0.9 | 293 | 86,489 | 125,310 | 219,688 | 77,054 | 9,710 | 518,416 |
| Biên Hòa | 9,028 | 3,282 | 1,775 | 14,084 | 1.6 | 530 | 156,346 | 226,524 | 397,131 | 139,290 | 35,399 | 959,264 |
| Nha Trang | 2,573 | 935 | 506 | 4,014 | 0.5 | 151 | 44,558 | 64,558 | 113,180 | 39,697 | 14,691 | 289,268 |
| Vũng Tàu | 2,933 | 1,066 | 577 | 4,576 | 0.5 | 172 | 50,794 | 73,594 | 129,021 | 45,253 | 9,283 | 317,141 |
| **Total** | **232,738** | **84,599** | **45,749** | **363,086** | **42** | **13,674** | **4,030,630** | **5,839,849** | **10,238,129** | **3,590,929** | **809,105** | **24,508,641** |

| **Supporting Table S6: Comparison of the projected number of hospitalized cases to Ministry of Health data** | | | | |
| --- | --- | --- | --- | --- |
|  | | **Projected number of hospitalized cases** | **The average number of reported hospitalized cases – Ministry of Health data (2016-2019)** | **Ratio of the projected to reported hospitalized cases** |
| Hồ Chí Minh | | 20,359 | 11,612 | 1.75 |
| Hà Nội | | 14,591 | 10,619 | 1.37 |
| Đà Nẵng | | 1,554 | 4,231 | 0.37 |
| Cần Thơ | | 2,440 | 954 | 2.56 |
| Thuận An | | 1,651 | 2,110 | 0.78 |
| Dĩ An | | 1,315 | 1,516 | 0.87 |
| Thủ Dầu Một | | 982 | 1,504 | 0.65 |
| Biên Hòa | | 1,775 | 1,757 | 1.01 |
| Nha Trang | | 506 | 1,568 | 0.32 |
| Vũng Tàu | | 577 | 884 | 0.65 |
| **Total** |  | 45,749 | 36,754 | 1.24 |

| **Supporting Table S7: Base case projected total cost and impact of the** ***Wolbachia* deployments (2020 US$ prices)** | | | | | | | | | | |
| --- | --- | --- | --- | --- | --- | --- | --- | --- | --- | --- |
| **Setting** | **Total cost of the intervention (US$)** | **DALYs averted** | **Averted cases that sought no formal treatment** | **Averted outpatient cases** | **Averted hospitalized cases averted** | **Total number of cases averted** | **Total cost of illness averted (US$)^1^** | **Total economic burden averted (US$)^2^** | **Breakeven year (from the start of the intervention)** | **Societal benefit-cost ratio** |
| Hồ Chí Minh | 76,002,063 | 68,215 | 1,776,452 | 645,730 | 349,193 | 2,771,698 | 129,184,363 | 132,125,986 | 13 | 1.74 |
| Hà Nội | 53,044,085 | 48,888 | 1,273,122 | 462,773 | 250,255 | 1,986,380 | 92,581,972 | 96,053,767 | 13 | 1.81 |
| Đà Nẵng | 9,003,297 | 5,205 | 135,552 | 49,272 | 26,645 | 211,494 | 9,857,396 | 10,377,131 | 19 | 1.15 |
| Cần Thơ | 8,420,004 | 8,176 | 212,910 | 77,392 | 41,851 | 332,191 | 15,482,906 | 15,922,973 | 12 | 1.89 |
| Thuận An | 4,402,016 | 5,533 | 144,084 | 52,374 | 28,322 | 224,807 | 10,477,887 | 10,676,213 | 10 | 2.43 |
| Dĩ An | 3,804,998 | 4,407 | 114,767 | 41,717 | 22,560 | 179,065 | 8,345,934 | 8,503,830 | 11 | 2.23 |
| Thủ Dầu Một | 2,338,347 | 3,289 | 85,656 | 31,135 | 16,837 | 133,644 | 6,228,921 | 6,335,899 | 9 | 2.71 |
| Biên Hòa | 8,524,624 | 5,946 | 154,840 | 56,284 | 30,437 | 241,588 | 11,260,043 | 11,693,085 | 16 | 1.37 |
| Nha Trang | 3,537,894 | 1,695 | 44,129 | 16,041 | 8,674 | 68,851 | 3,209,056 | 3,504,532 | - | 0.99 |
| Vũng Tàu | 2,235,582 | 1,932 | 50,305 | 18,286 | 9,888 | 78,488 | 3,658,206 | 3,858,388 | 13 | 1.73 |
| **Total** | **171,312,910** | **153,285** | **3,991,817** | **1,451,004** | **784,663** | **6,228,208** | **290,286,683** | **299,051,805** | **-** | **1.75** |
| *^1^The cost of illness is the cost a speciﬁc disease or condition imposes on society (i.e the direct costs and productivity costs associated with dengue cases).*  *^2^The economic burden includes the cost of illness but also the costs associated with government’s current dengue prevention and control activities.* | | | | | | | | | | |

| **Supporting Table S8: Base case setting specific cost-effectiveness ratios (2020 US$ prices)** | | | | | | |
| --- | --- | --- | --- | --- | --- | --- |
| **Setting** | **Projected incidence per 100,000 population^1^** | **Gross cost-effectiveness ratio** | **Incremental cost-effectiveness ratio - health care provider perspective** | **Incremental cost-effectiveness ratio - health sector perspective** | **Incremental cost-effectiveness ratio - societal perspective** | **Incremental cost-effectiveness ratio - societal perspective (excluding the productivity gains related to prevented excess mortality)** |
| Hồ Chí Minh | 1,797 | 1,114 | 776 | 430 | -780 | -493 |
| Hà Nội | 1,438 | 1,085 | 719 | 373 | -809 | -522 |
| Đà Nẵng | 1,087 | 1,730 | 1,335 | 989 | -164 | 123 |
| Cần Thơ | 1,568 | 1,030 | 681 | 335 | -864 | -577 |
| Thuận An | 2,198 | 796 | 465 | 119 | -1,098 | -811 |
| Dĩ An | 2,199 | 863 | 533 | 187 | -1,030 | -743 |
| Thủ Dầu Một | 2,423 | 711 | 383 | 38 | -1,183 | -896 |
| Biên Hòa | 1,334 | 1,434 | 1,066 | 720 | -460 | -173 |
| Nha Trang | 950 | 2,088 | 1,618 | 1,273 | 194 | 481 |
| Vũng Tàu | 1,281 | 1,157 | 759 | 413 | -736 | -450 |
| **Overall** | **1,627** | **1,118** | **708** | **420** | **-776** | **-546** |
| *^1^ Based on the projected case numbers and the total population within the administrative district boundary (Table 2).* | | | | | | |

| **Supporting Table S9: Values within the sensitivity analysis - (2020 US$ prices)** | | | | | | | | | | | | | | | | |
| --- | --- | --- | --- | --- | --- | --- | --- | --- | --- | --- | --- | --- | --- | --- | --- | --- |
|  | **Base case results** | **Using the confidence interval on the baseline incidence of dengue cases** | | **Assuming an alternative breakdown of the type of care dengue cases receive** | **DALY weights - accounting for only acute symptoms** | **Using the confidence intervals for the cost of illness of non-fatal cases** | | **Varying the assumed annual growth in case numbers (0-5%)** | | **Varying the assumed effectiveness of the Wolbachia deployments (65%-85%)** | | **Varying the assumed duration of the benefits (10-25 years)** | | **Assuming the long-term monitoring phase lasts 20 years** | **Using a 0% discount rate for the health effects** | **Using a 6% discount rate for the costs** |
|  |  | **Lower** | **Higher** |  |  | **Lower** | **Higher** | **Lower** | **Higher** | **Lower** | **Higher** | **Lower** | **Higher** |  |  |  |
| Baseline DALY burden | 13,674 | 11,454 | 16,627 | 13,745 | 6,417 | 13,674 | 13,674 | 13,674 | 13,674 | 13,674 | 13,674 | 13,674 | 13,674 | 13,674 | 13,674 | 13,674 |
| Number of DALYs averted (thousands) | 153 | 128 | 186 | 154 | 64 | 153 | 153 | 136 | 243 | 133 | 174 | 84 | 184 | 153 | 153 | 153 |
| Total cost of the intervention (million) | 171 | 171 | 171 | 171 | 171.31 | 171 | 171 | 171 | 171 | 171 | 171 | 171 | 171 | 120 | 125 | 174 |
| Cost per person reached | 8.56 | 8.56 | 8.56 | 8.56 | 8.56 | 8.56 | 8.56 | 8.56 | 8.56 | 8.56 | 8.56 | 8.56 | 8.56 | 5.99 | 6.25 | 8.68 |
| Societal benefit-cost ratio | 1.75 | 1.47 | 2.11 | 2.69 | 1.75 | 1.42 | 2.16 | 1.55 | 2.74 | 1.52 | 1.97 | 0.95 | 2.09 | 2.49 | 2.39 | 1.72 |
| Gross cost-effectiveness ratio | 1,118 | 1,334 | 919 | 1,111 | 2,660 | 1,118 | 1,118 | 1,264 | 705 | 1,290 | 986 | 2,050 | 933 | 783 | 816 | 1,133 |
| ICER - health sector perspective | 420 | 625 | 231 | -45 | 999 | 557 | 263 | 558 | 28 | 583 | 295 | 1349 | 239 | 85 | 118 | 435 |
| ICER - societal perspective (excluding the productivity gains related to prevented excess mortality) | -546 | -341 | -735 | -1,592 | -1,301 | -177 | -1,013 | -408 | -938 | -383 | -671 | 383 | -727 | -881 | -848 | -531 |
| *See Table 5 for parameter ranges. WMP; World Mosquito Program, ICER; Incremental cost-effectiveness ratio.*  *Negative ratios (“Cost savings”) in the case indicate that the economic benefits of the health intervention relative to the comparator outweighed the cost of the intervention. Note that these “Cost savings” include non-fiscal costs* | | | | | | | | | | | | | | | | |

| **Supporting Table S10: Setting specific cost-effectiveness ratios – when assuming only 10 years of benefits (2020 US$ prices)** | | | | | | | |
| --- | --- | --- | --- | --- | --- | --- | --- |
| **Setting** | **Gross cost-effectiveness ratio** | | **Incremental cost-effectiveness ratio - health care provider perspective** | **Incremental cost-effectiveness ratio - health sector perspective** | | **Incremental cost-effectiveness ratio - societal perspective** | **Incremental cost-effectiveness ratio - societal perspective (excluding the productivity gains related to prevented excess mortality)** |
| Hồ Chí Minh | | 2,044 | 1,704 | | 1,358 | 150 | 437 |
| Hà Nội | | 1,990 | 1,621 | | 1,275 | 97 | 384 |
| Đà Nẵng | | 3,173 | 2,773 | | 2,427 | 1,279 | 1,566 |
| Cần Thơ | | 1,889 | 1,538 | | 1,192 | -4 | 282 |
| Thuận An | | 1,460 | 1,127 | | 781 | -434 | -147 |
| Dĩ An | | 1,584 | 1,251 | | 905 | -310 | -23 |
| Thủ Dầu Một | | 1,304 | 975 | | 629 | -590 | -303 |
| Biên Hòa | | 2,630 | 2,259 | | 1,913 | 736 | 1,023 |
| Nha Trang | | 3,830 | 3,352 | | 3,006 | 1,936 | 2,223 |
| Vũng Tàu | | 2,123 | 1,719 | | 1,373 | 229 | 516 |
| **Overall** | | **2,050** | **1,635** | | **1,349** | **156** | **383** |
| *^1^ Based on the projected case numbers and the total population within the administrative district boundary (Table 2).* | | | | | | | |

| **Supporting Table S11: The cost-effectiveness and cost benefit of the Wolbachia deployments for different intervention costs** | | | | | | | | |
| --- | --- | --- | --- | --- | --- | --- | --- | --- |
|  | **Average discounted cost per person covered (US$)** | | | | | | | |
|  | **US$3** | **US$5** | **US$8.56**  **(Base case)** | **US$12.00** | **US$3** | **US$5** | **US$8.56**  **(Base case)** | **US$12.00** |
|  | **10 years of benefits** | | | | **20 years of benefits** | | | |
| Total cost | 60,059,504 | 100,099,173 | 171,369,785 | 240,238,016 | 60,059,504 | 100,099,173 | 171,369,785 | 240,238,016 |
| Gross cost-effectiveness ratio | 719 | 1,198 | 2,051 | 2,875 | 392 | 653 | 1,118 | 1,567 |
| Incremental cost-effectiveness ratio - health care provider perspective | 303 | 783 | 1,636 | 2,460 | -18 | 244 | 709 | 1,158 |
| Incremental cost-effectiveness ratio - health sector perspective | 18 | 497 | 1,350 | 2,174 | -306 | -45 | 420 | 869 |
| Incremental cost-effectiveness ratio - societal perspective | “Cost saving” (-1,175) | “Cost saving” (-696) | 157 | 981 | “Cost saving” (-1,502) | “Cost saving” (-1,241) | “Cost saving” (-776) | “Cost saving” (-327) |
| Incremental cost-effectiveness ratio - societal perspective (excluding the productivity gains related to prevented excess mortality) | “Cost saving” (-948) | “Cost saving” (-469) | 384 | 1,208 | “Cost saving (-1,272) | “Cost saving” (-1,011) | “Cost saving” (-546) | “Cost saving” (-97) |
| Societal benefit-cost ratio | 2.72 | 1.63 | 0.95 | 0.68 | 4.98 | 2.99 | 1.75 | 1.24 |
| *Negative ratios (“Cost savings”) in the case indicate that the economic benefits of the health intervention relative to the comparator outweighed the cost of the intervention. Note that these “Cost savings” include non-fiscal costs.* | | | | | | | | |

| **The CHEERS 2022 checklist** | | | |
| --- | --- | --- | --- |
| Section/topic | Item No | Guidance for reporting | Reported in section |
| **Title** | | |  |
| Title | 1 | Identify the study as an economic evaluation and specify the interventions being compared. | Title |
| **Abstract** | | |  |
| Abstract | 2 | Provide a structured summary that highlights context, key methods, results, and alternative analyses. | Abstract |
| **Introduction** | | |  |
| Background and objectives | 3 | Give the context for the study, the study question, and its practical relevance for decision making in policy or practice. | Introduction |
| **Methods** | | |  |
| Health economic analysis plan | 4 | Indicate whether a health economic analysis plan was developed and where available. | Supporting information: Further methodological details |
| Study population | 5 | Describe characteristics of the study population (such as age range, demographics, socioeconomic, or clinical characteristics). | Methods: The selected settings and dengue incidence and the Supporting information: Project sites and stratification of dengue cases |
| Setting and location | 6 | Provide relevant contextual information that may influence findings. | Methods: The selected settings and dengue incidence and Table 1 |
| Comparators | 7 | Describe the interventions or strategies being compared and why chosen. | Methods: The economic evaluation |
| Perspective | 8 | State the perspective(s) adopted by the study and why chosen. | Methods: The economic evaluation and Box 1 |
| Time horizon | 9 | State the time horizon for the study and why appropriate. | Methods: The economic evaluation |
| Discount rate | 10 | Report the discount rate(s) and reason chosen. | Methods: The economic evaluation |
| Selection of outcomes | 11 | Describe what outcomes were used as the measure(s) of benefit(s) and harm(s). | Methods: Effectiveness of the Wolbachia deployments, Box 1 and Supporting information |
| Measurement of outcomes | 12 | Describe how outcomes used to capture benefit(s) and harm(s) were measured. | Methods: Effectiveness of the Wolbachia deployments, Box 1 and Supporting information |
| Valuation of outcomes | 13 | Describe the population and methods used to measure and value outcomes. | Methods: Effectiveness of the Wolbachia deployments, Methods: Health burden and economic burden of dengue and Supporting information |
| Measurement and valuation of resources and costs | 14 | Describe how costs were valued. | Methods: Costs of the Wolbachia deployments and Supporting information |
| Currency, price date, and conversion | 15 | Report the dates of the estimated resource quantities and unit costs, plus the currency and year of conversion. | Methods: The economic evaluation |
| Rationale and description of model | 16 | If modelling is used, describe in detail and why used. Report if the model is publicly available and where it can be accessed. | Methods: The economic evaluation |
| Analytics and assumptions | 17 | Describe any methods for analysing or statistically transforming data, any extrapolation methods, and approaches for validating any model used. | NA |
| Characterising heterogeneity | 18 | Describe any methods used for estimating how the results of the study vary for subgroups. | Supporting information: Further methodological details |
| Characterising distributional effects | 19 | Describe how impacts are distributed across different individuals or adjustments made to reflect priority populations. | Supporting information: Further methodological details |
| Characterising uncertainty | 20 | Describe methods to characterise any sources of uncertainty in the analysis. | Methods: The economic evaluation and Table 3 |
| Approach to engagement with patients and others affected by the study | 21 | Describe any approaches to engage patients or service recipients, the general public, communities, or stakeholders (such as clinicians or payers) in the design of the study. | Supporting information |
| **Results** | | |  |
| Study parameters | 22 | Report all analytic inputs (such as values, ranges, references) including uncertainty or distributional assumptions. | Table 2 and Supporting information |
| Summary of main results | 23 | Report the mean values for the main categories of costs and outcomes of interest and summarise them in the most appropriate overall measure. | Table 3 and results |
| Effect of uncertainty | 24 | Describe how uncertainty about analytic judgments, inputs, or projections affect findings. Report the effect of choice of discount rate and time horizon, if applicable. | Results: Sensitivity analysis |
| Effect of engagement with patients and others affected by the study | 25 | Report on any difference patient/service recipient, general public, community, or stakeholder involvement made to the approach or findings of the study | NA |
| **Discussion** | | |  |
| Study findings, limitations, generalisability, and current knowledge | 26 | Report key findings, limitations, ethical or equity considerations not captured, and how these could affect patients, policy, or practice. | Discussion and Supporting information |
| **Other relevant information** | | | |
| Source of funding | 27 | Describe how the study was funded and any role of the funder in the identification, design, conduct, and reporting of the analysis | Funding statement |
| Conflicts of interest | 28 | Report authors conflicts of interest according to journal or International Committee of Medical Journal Editors requirements. | Conflicts of interest statement |

8.	The World Bank. GDP per capita (current US$). Available at: <https://data.worldbank.org/indicator/NY.GDP.PCAP.CD>.

9.	Tan-Torres Edejer T, Baltussen R, Adam Ta, et al. Making choices in health: WHO guide to cost-effectiveness analysis. **2003**.

10.	World Bank. GDP deflator (base year varies by country). Available at: <https://data.worldbank.org/indicator/NY.GDP.DEFL.ZS>.

11.	Turner HC, Lauer JA, Tran BX, Teerawattananon Y, Jit M. Adjusting for Inflation and Currency Changes Within Health Economic Studies. Value Health **2019**; 22(9): 1026-32.
